# Sleep Irregularity and Risk of Incident Hypertension in the UK Biobank

**DOI:** 10.64898/2026.11.12.26367978

**Authors:** Kaitlin S. Potts, Sina Kianersi, Tariq Faquih, Raymond Noordam, Martin K. Rutter, Tamar Sofer, Sean Cain, Daniel Windred, Angus C. Burns, Andrew J.K. Phillips, Susan Redline, Heming Wang, Tianyi Huang

**Author notes:** These authors jointly supervised this work (Heming Wang, Tianyi Huang).

## Abstract

**Background:** Irregular sleep patterns are common and have been associated with higher cardiovascular disease (CVD) risk. Hypertension is the leading modifiable cause of CVD, but evidence linking irregular sleep to hypertension is inconsistent. The objective of this study was to quantify associations between irregular sleep and incident hypertension.

**Methods:** We conducted a prospective cohort study among 55,572 UK Biobank participants without hypertension or anti-hypertensive medication use who provided ≥5 valid days of accelerometry in 2013- 2015 (60% women, mean age 61 years). We measured sleep regularity using the intra-individual standard deviation (SD) of sleep duration and the sleep regularity index (SRI), a measure of the similarity of sleep-wake patterns from one day to the next. Cox proportional hazards models estimated adjusted hazard ratios (HR) for incident hypertension, identified through linked health records, by levels of sleep regularity adjusting for demographic, socioeconomic, and lifestyle factors as well as employment schedule. We also evaluated differences by demographic, clinical, and sleep-related (defined by duration, efficiency, timing, and sleep disorders) subgroups, and interaction with genetic susceptibility to hypertension.

**Results:** During 7.9 years of follow-up, 5,713 participants developed hypertension. Participants with sleep duration SD >90 minutes had an 18% higher risk of incident hypertension compared to those with sleep duration SD ≤30 minutes (HR: 1.18; 95% confidence interval (CI): 1.06, 1.31). Similarly, participants in the lowest SRI quintile had a 40% higher risk than those in the highest SRI quintile (HR: 1.40; 95% CI: 1.29, 1.52). These associations were generally consistent across demographic, health, and sleep-defined subgroups. Sleep irregularity and genetic susceptibility contributed independently to hypertension risk, without evidence of interaction.

**Conclusions:** Irregular sleep patterns were associated with higher incident hypertension risk independent of established risk factors and genetic susceptibility. Maintaining regular sleep schedules may represent a modifiable behavioral target for hypertension prevention.

**Clinical Perspective:** *What is new?:*

- In this large prospective study using objectively measured sleep from wrist accelerometry, greater overall variability in sleep duration and more irregular day-to-day sleep–wake patterns were independently associated with higher risk of incident hypertension.
- Associations were robust to adjustment for potential confounding by demographic, socioeconomic, clinical, and lifestyle factors and were generally consistent across demographic, clinical, and sleep-defined subgroups.
- Sleep regularity and genetic susceptibility to hypertension were independently associated with hypertension risk, with no evidence that inherited risk modified the association between irregular sleep and incident hypertension.

*What are the clinical implications?:*

- Maintaining consistent sleep schedules may represent an important modifiable behavioral target for hypertension prevention alongside established lifestyle recommendations.
- The association between irregular sleep and hypertension was observed regardless of genetic susceptibility, suggesting that improving sleep regularity may benefit individuals across the spectrum of inherited hypertension risk.
- These findings support future randomized clinical trials to determine whether interventions that improve sleep regularity can reduce blood pressure and prevent hypertension.

## Introduction

Hypertension is the most prevalent modifiable risk factor for cardiovascular disease (CVD), affecting more 1.4 billion, or a third, of adults aged 30-79 years worldwide^1^. Despite effective pharmacologic treatments, prevention remains a major public health priority given the high prevalence of hypertension, its strong contribution to cardiovascular morbidity and mortality, and its difficulty to control after it develops^2^. While behavioral strategies have traditionally focused on diet and physical activity, sleep is increasingly recognized as a core behavioral component of cardiovascular health^3^.

Sleep has traditionally been characterized by duration and quality, but emerging evidence highlights sleep regularity as a distinct and clinically relevant dimension of sleep^4^. Recent evidence suggests that maintaining a consistent sleep schedule may have even greater health benefits compared to attaining adequate sleep duration, and that the benefits of regular sleep are independent of sleep duration^5,6^. Irregular sleep patterns are common in modern societies and may contribute to circadian disruption or misalignment, which are implicated in adverse cardiometabolic outcomes^7^. While several cross-sectional and prospective observational studies have linked irregular sleep to both CVD risk factors and outcomes^5,8,9^, reviews have highlighted that evidence for an association between sleep regularity and hypertension remains inconsistent^10–12^; Associations may also vary according to other clinical factors, including average sleep duration and the presence of obstructive sleep apnea (OSA) or prediabetes^11,13–15^. Prior studies investigating the association between irregular sleep and hypertension have been limited by cross-sectional designs or, among prospective studies, reliance on self-reported sleep regularity, small sample sizes due in part to high baseline hypertension prevalence, short follow-up and few incident cases, or inadequate participant characterization for confounder control. In addition, few studies have examined complementary accelerometry-derived sleep regularity metrics, limiting insight into whether different aspects of irregular sleep confer similar risks.

With strong evidence of heritability, genetic susceptibility to hypertension, as captured by polygenic risk score (PRS), has been shown to have varying effects across different groups (e.g., by sex and obesity status)^16^. While interactions between short or long sleep duration and blood pressure genetic loci have been reported^17^, the interaction between irregular sleep schedules and genetic susceptibility to hypertension remains unexplored. Whether the association between irregular sleep patterns and hypertension risk differs by level of hypertension PRS remains largely unexplored and this information may inform precision prevention by identifying individuals who may be most likely to benefit from improving sleep regularity.

To address these gaps, we examined associations of two complementary measures of sleep irregularity with risk for incident hypertension in the UK Biobank (UKB), a large prospective cohort with accelerometer-derived sleep data and long-term follow-up via linkage to healthcare records. We also assessed potential effect modification by demographic, clinical, and other sleep factors, and interaction by genetic susceptibility for hypertension. We hypothesized that greater sleep irregularity would be associated with higher risk of incident hypertension.

## Methods

### Sample and study design

The UKB is a prospective cohort study that enrolled over 502,000 United Kingdom residents aged 40-69 years at the time of the baseline visit (2006-2010)^18^. Baseline assessments included questionnaires, interviews, physical measurements, and biologic sample collection to capture a broad range of socio-demographic, lifestyle, and health characteristics. The study was approved by the NHS North-West Multi-Centre Research Ethics Committee, and all participants gave informed electronic consent. These data are available with an approved UKB application, and this study was conducted under UKB application ID 85501.

Between 2013 and 2015, a random sample of 236,519 UKB participants with email addresses were invited to participate in an accelerometer study^19^ of whom 103,610 provided up to 7 days of data, using Axivity AX3 triaxial devices worn on the dominant wrist (Supplementary Figure S1 for participant flowchart). We excluded individuals with less than five nights of valid accelerometry data, following methods described previously to identify valid recordings^20^, missing or extreme (>4 hours) sleep duration SD, or missing sleep regularity index (SRI) (n=23,976 excluded). Finally, we excluded 24,062 participants with likely prevalent hypertension any time prior to accelerometry, identified through medical records, self-report diagnoses, self-reported anti-hypertensive medication use, or anti-hypertensive medications recorded during nurse interviews, based on a published UKB antihypertensive medications list^21^.

### Sleep measures

Raw accelerometer data were processed by R package *GGIR* (version 1.5.12) as described by Jones et al.^20^. Briefly, 100-Hz triaxial acceleration signals were summarized to 5-second epochs, classified as sleep or wake, and used to identify the main sleep period time (SPT) window. The GGIR sleep detection algorithm has been validated against polysomnography for detecting the main sleep period even in the absence of diaries^22^. Using one SPT window per 24-hour period, *GGIR* estimated average sleep metrics across all available nights per participant, including sleep duration, sleep duration SD (the intra-individual standard deviation of sleep duration across all nights), sleep maintenance efficiency (percentage time asleep in SPT window), and sleep midpoint timing. We additionally calculated the sleep regularity index (SRI) using the *sleepreg* package as described previously^6,23^. The SRI is based on the probability of being in the same sleep-wake state at any two time points 24-h apart, averaged across the recording^24,25^. Briefly, the SRI used here is based on one-minute sleep/wake epochs including classification of naps and nighttime awakenings, and nights were excluded if their GGIR estimated SPT window was considered invalid based on the criteria described by Winded et al.^23^. A minimum of five valid nights were required. SRI Values closer to 100 reflect more regular sleep–wake patterns, whereas values closer to 0 represent increasingly random patterns. We focused on both sleep duration SD and SRI to capture complementary dimensions of sleep irregularity. Sleep duration SD reflects night-to-night variability in sleep duration and may be more clinically intuitive, while the SRI measures sleep-wake pattern regularity across consecutive days, capturing variability in duration, timing, nighttime wake episodes, and naps, potentially making it more sensitive to circadian disruption and irregularly fragmented sleep^25^. The SRI is slightly more sensitive to variation in timing compared to variation in duration and operates on a consecutive timescale, while sleep duration SD is agnostic to the ordering of nights and quantifies variation in duration only, not timing. Higher sleep duration SD and lower SRI each indicate more irregular patterns.

### Identification of hypertension and genetic risk

Incident hypertension occurring between the end date of accelerometry and the end of follow- up was identified using the UKB “first occurrence” algorithm to capture the earliest date of 3-character International Classification of Diseases, 10th Revision (ICD10) code I10, essential (primary) hypertension. The first occurrence algorithms incorporate linked medical records from the National Health Service covering hospital inpatient data, coded primary care data, and cancer and death registries, as well as interview-verified self-report diagnoses from study visits^26^. Hypertension identified through these UKB sources has been associated with expected hypertensive cardiac remodeling, suggesting validity^21^.

Genetic predisposition to hypertension was measured using both a polygenic risk score (PRS) for hypertension and self-reported history of hypertension among participants’ parents or siblings (family history). We used the Standard PRS described by Thompson et al., which was not trained on any UKB data^27^. Analyses using the PRS were restricted to participants self-reporting White/European ethnic background.

### Statistical analysis

Sleep duration SD and SRI were modeled categorically for primary analysis, consistent with previous research. Sleep duration SD was categorized as ≤30, 31-45, 46-60, 61-90, and >90 minutes^28–30^ to facilitate clinical interpretation, and SRI was categorized into quintiles^6^. About 9% of the sample had sleep duration SD >90 minutes; we used quintiles of sleep duration SD in a sensitivity analysis to aid comparison of results to those found using SRI quintiles. We also evaluated sleep regularity continuously (per SD of each metric) and as binary variables, comparing the most irregular sleepers (>90 minutes sleep duration SD and 0-20^th^ SRI percentiles) to all others. Using the continuous sleep duration SD and SRI, potential nonlinearity was further evaluated using natural cubic splines with knots placed at the 10^th^, 50^th^, and 90^th^ percentiles^31^. Likelihood ratio tests comparing spline and linear models evaluated statistical significance of nonlinearity. End of follow-up was October 31, 2022, corresponding to the censoring date for England hospital data available in the UKB data used for this analysis (version 18.1, available from March 2025; the majority of hospital data came from England but from Scotland and Wales data were also used and their corresponding end of follow-up dates were within 5 months of the date used). Follow-up time was calculated from the end date of accelerometer recording to the earliest occurrence of incident hypertension, death, loss to follow-up, or end of follow-up, whichever came first. Hazard ratios (HRs) and 95% confidence intervals (CIs) for incident hypertension by level of sleep regularity were estimated with Cox proportional hazards regression. The proportional hazards assumption was evaluated using Schoenfeld residuals.

We estimated associations using sequentially adjusted models selected *a priori* based on known or plausible associations with sleep regularity and hypertension to control for confounding. Supplementary Table S1. provides UKB data fields, source questions, and coding descriptions for all variables used. Age and sex were included in the minimally adjusted model (Model 0). Model 1 additionally adjusted for self-reported ethnic background, Townsend deprivation index, employment/shiftwork, and family history of hypertension. Model 2, used for primary interpretation, further adjusted for lifestyle factors including smoking history, alcohol use, physical activity, and healthy diet score. In a sensitivity model we additionally included health factors that may be possible mediators or colliders, including body mass index (BMI), diabetes, use of cholesterol-lowering medication, and presence of depression or anxiety symptoms. In a second sensitivity model we also adjusted for baseline self-reported chronotype, sleepiness, and insomnia, diagnosed sleep disorders any time until the accelerometer visit (ICD-10 code G47), and accelerometer-measured average sleep duration. Missing covariate data were minimal (most <1%; highest was 3% missing on depression/anxiety, used only in sensitivity model), so we applied single imputation using medians and modes for continuous and categorical variables, respectively (Supplementary Table S2 show numbers imputed per variable).

We further evaluated associations within pre-specified demographic, health, and sleep-related subgroups using Model 2 and binary sleep regularity variables to examine consistency of associations. Stratified analyses were conducted by age at accelerometry, sex, ethnicity, family history of hypertension, hypertension PRS (low risk: tertile 1, medium risk: tertile 2, high risk: tertile 3), baseline BMI, baseline self-reported sleep characteristics (insomnia symptoms, excessive sleepiness, snoring, and chronotype), and health conditions identified any time prior to accelerometry (diabetes history and diagnosed sleep disorders). We also stratified by other accelerometry-derived sleep characteristics including sleep duration (short: <20^th^ percentile, intermediate: 20^th^ to <80^th^ percentile, long: ≥80^th^ percentile), sleep midpoint time (early: <20^th^ percentile, intermediate: 20^th^ to <80^th^ percentile, late: ≥80^th^ percentile), and sleep efficiency (inefficient: <0.8, efficient: ≥0.8). We reported interaction p-values from Wald tests for the product term between binary sleep regularity variables and the subgroups; these analyses were considered exploratory.

To evaluate interaction with genetic susceptibility, we estimated joint HRs for sleep regularity (using binary variables) and genetic risk (PRS and family history) using a common low-risk reference group (regular sleep and low-medium genetic risk), following the approach of VanderWeele and Knol^32^. We assessed multiplicative and additive interaction, with additive interaction quantified by the relative excess risk due to interaction (RERI). Ninety-five percent confidence intervals were estimated using the variance recovery method^33,34^.

Several sensitivity analyses were performed to interrogate robustness of results. First, we excluded individuals with high blood pressure readings at the UKB baseline visit (SBP>130 or DBP>80) who were not otherwise excluded based on the criteria of prevalent hypertension described above. Second, we repeated the analysis censoring at March 23, 2020, the start date of the COVID-19 lockdown in the UK, to account for disruptions to healthcare during and after the pandemic that may have delayed hypertension diagnoses. Third, we excluded those diagnosed with hypertension in the first year of follow-up in case they represented delayed diagnoses or reverse causation. Finally, because sleep duration SD was heavily right-skewed, we repeated the analysis excluding those with sleep duration SD >2 hours.

## Results

Among the 55,572 individuals included in analyses, 60% were female and 97% self-reported as White and European ethnic background (Table 1). The mean (SD) age at the time of accelerometry was 61(8) years. At the UKB baseline visit, which occurred a median of 5.7 years earlier than the accelerometry assessment, 66% were employed (12% of those employed worked in shift jobs), 7% were current smokers, 33% had depressive or anxiety symptoms, and the mean BMI was 26 kg/m^2^. Almost 90% of the sample provided 7 valid nights of data (only 2% had 5 nights).

**Table 1.**
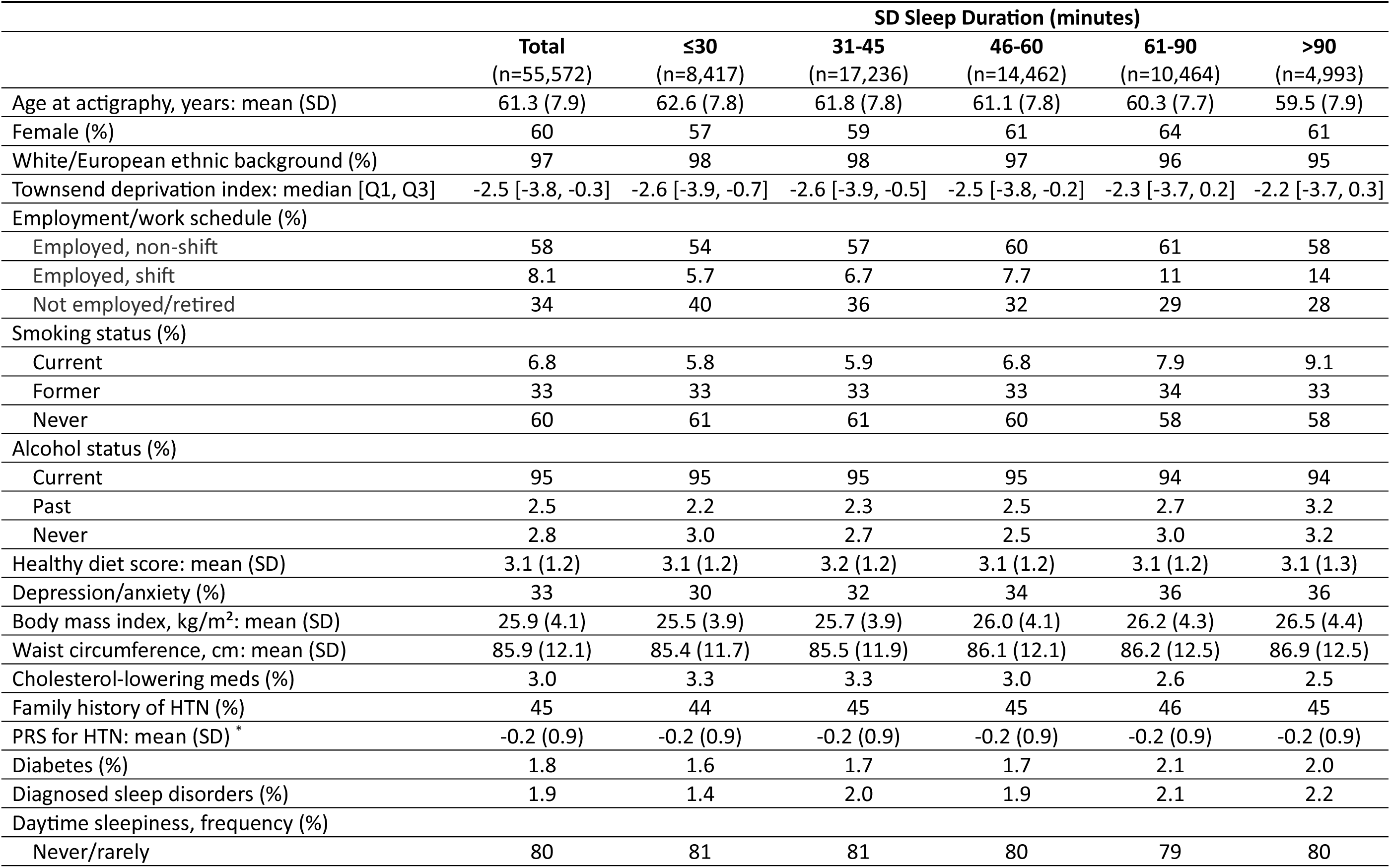
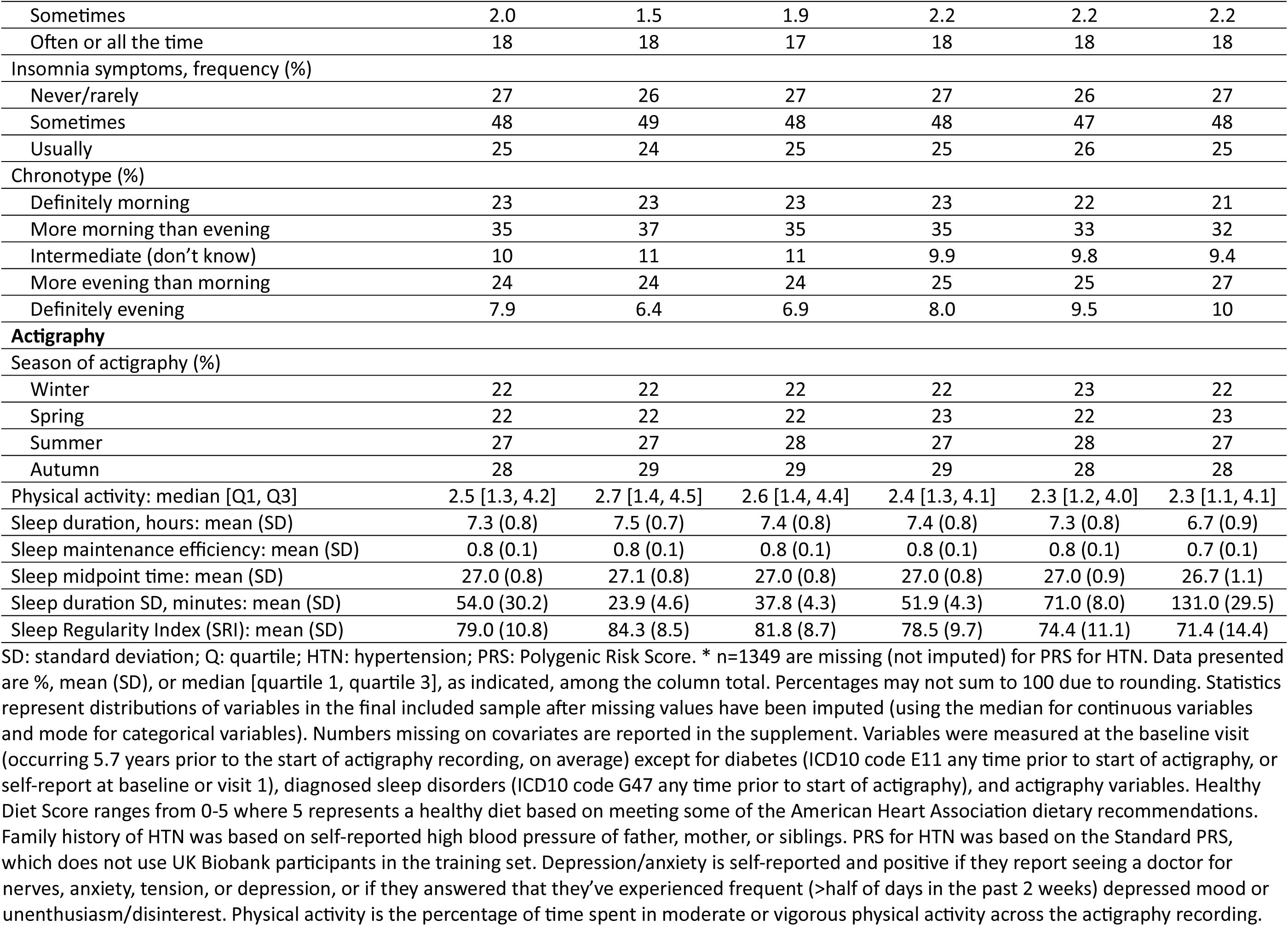
Description of participants by level of sleep regularity (SD sleep duration).

|  | **Total** | **SD Sleep Duration (minutes)** |  |  |  |  |
|---|---|---|---|---|---|---|
|  | **Total**<br>(n=55,572) | **≤30**<br>(n=8,417) | **31-45**<br>(n=17,236) | **46-60**<br>(n=14,462) | **61-90**<br>(n=10,464) | **>90**<br>(n=4,993) |
| Age at actigraphy, years: mean (SD) | 61.3 (7.9) | 62.6 (7.8) | 61.8 (7.8) | 61.1 (7.8) | 60.3 (7.7) | 59.5 (7.9) |
| Female (%) | 60 | 57 | 59 | 61 | 64 | 61 |
| White/European ethnic background (%) | 97 | 98 | 98 | 97 | 96 | 95 |
| Townsend deprivation index: median [Q1, Q3] | -2.5 [-3.8, -0.3] | -2.6 [-3.9, -0.7] | -2.6 [-3.9, -0.5] | -2.5 [-3.8, -0.2] | -2.3 [-3.7, 0.2] | -2.2 [-3.7, 0.3] |
| Employment/work schedule (%) |  |  |  |  |  |  |
| Employed, non-shift | 58 | 54 | 57 | 60 | 61 | 58 |
| Employed, shift | 8.1 | 5.7 | 6.7 | 7.7 | 11 | 14 |
| Not employed/retired | 34 | 40 | 36 | 32 | 29 | 28 |
| Smoking status (%) |  |  |  |  |  |  |
| Current | 6.8 | 5.8 | 5.9 | 6.8 | 7.9 | 9.1 |
| Former | 33 | 33 | 33 | 33 | 34 | 33 |
| Never | 60 | 61 | 61 | 60 | 58 | 58 |
| Alcohol status (%) |  |  |  |  |  |  |
| Current | 95 | 95 | 95 | 95 | 94 | 94 |
| Past | 2.5 | 2.2 | 2.3 | 2.5 | 2.7 | 3.2 |
| Never | 2.8 | 3.0 | 2.7 | 2.5 | 3.0 | 3.2 |
| Healthy diet score: mean (SD) | 3.1 (1.2) | 3.1 (1.2) | 3.2 (1.2) | 3.1 (1.2) | 3.1 (1.2) | 3.1 (1.3) |
| Depression/anxiety (%) | 33 | 30 | 32 | 34 | 36 | 36 |
| Body mass index, kg/m<sup>2</sup>: mean (SD) | 25.9 (4.1) | 25.5 (3.9) | 25.7 (3.9) | 26.0 (4.1) | 26.2 (4.3) | 26.5 (4.4) |
| Waist circumference, cm: mean (SD) | 85.9 (12.1) | 85.4 (11.7) | 85.5 (11.9) | 86.1 (12.1) | 86.2 (12.5) | 86.9 (12.5) |
| Cholesterol-lowering meds (%) | 3.0 | 3.3 | 3.3 | 3.0 | 2.6 | 2.5 |
| Family history of HTN (%) | 45 | 44 | 45 | 45 | 46 | 45 |
| PRS for HTN: mean (SD)<sup>*</sup> | -0.2 (0.9) | -0.2 (0.9) | -0.2 (0.9) | -0.2 (0.9) | -0.2 (0.9) | -0.2 (0.9) |
| Diabetes (%) | 1.8 | 1.6 | 1.7 | 1.7 | 2.1 | 2.0 |
| Diagnosed sleep disorders (%) | 1.9 | 1.4 | 2.0 | 1.9 | 2.1 | 2.2 |
| Daytime sleepiness, frequency (%) |  |  |  |  |  |  |
| Never/rarely | 80 | 81 | 81 | 80 | 79 | 80 |
| Sometimes | 2.0 | 1.5 | 1.9 | 2.2 | 2.2 | 2.2 |
| Often or all the time | 18 | 18 | 17 | 18 | 18 | 18 |
| Insomnia symptoms, frequency (%) |  |  |  |  |  |  |
| Never/rarely | 27 | 26 | 27 | 27 | 26 | 27 |
| Sometimes | 48 | 49 | 48 | 48 | 47 | 48 |
| Usually | 25 | 24 | 25 | 25 | 26 | 25 |
| Chronotype (%) |  |  |  |  |  |  |
| Definitely morning | 23 | 23 | 23 | 23 | 22 | 21 |
| More morning than evening | 35 | 37 | 35 | 35 | 33 | 32 |
| Intermediate (don’t know) | 10 | 11 | 11 | 9.9 | 9.8 | 9.4 |
| More evening than morning | 24 | 24 | 24 | 25 | 25 | 27 |
| Definitely evening | 7.9 | 6.4 | 6.9 | 8.0 | 9.5 | 10 |
| **Actigraphy** |  |  |  |  |  |  |
| Season of actigraphy (%) |  |  |  |  |  |  |
| Winter | 22 | 22 | 22 | 22 | 23 | 22 |
| Spring | 22 | 22 | 22 | 23 | 22 | 23 |
| Summer | 27 | 27 | 28 | 27 | 28 | 27 |
| Autumn | 28 | 29 | 29 | 29 | 28 | 28 |
| Physical activity: median [Q1, Q3] | 2.5 [1.3, 4.2] | 2.7 [1.4, 4.5] | 2.6 [1.4, 4.4] | 2.4 [1.3, 4.1] | 2.3 [1.2, 4.0] | 2.3 [1.1, 4.1] |
| Sleep duration, hours: mean (SD) | 7.3 (0.8) | 7.5 (0.7) | 7.4 (0.8) | 7.4 (0.8) | 7.3 (0.8) | 6.7 (0.9) |
| Sleep maintenance efficiency: mean (SD) | 0.8 (0.1) | 0.8 (0.1) | 0.8 (0.1) | 0.8 (0.1) | 0.8 (0.1) | 0.7 (0.1) |
| Sleep midpoint time: mean (SD) | 27.0 (0.8) | 27.1 (0.8) | 27.0 (0.8) | 27.0 (0.8) | 27.0 (0.9) | 26.7 (1.1) |
| Sleep duration SD, minutes: mean (SD) | 54.0 (30.2) | 23.9 (4.6) | 37.8 (4.3) | 51.9 (4.3) | 71.0 (8.0) | 131.0 (29.5) |
| Sleep Regularity Index (SRI): mean (SD) | 79.0 (10.8) | 84.3 (8.5) | 81.8 (8.7) | 78.5 (9.7) | 74.4 (11.1) | 71.4 (14.4) |

SD: standard deviation; Q: quartile; HTN: hypertension; PRS: Polygenic Risk Score. \* n=1349 are missing (not imputed) for PRS for HTN. Data presented are %, mean (SD), or median [quartile 1, quartile 3], as indicated, among the column total. Percentages may not sum to 100 due to rounding. Statistics represent distributions of variables in the final included sample after missing values have been imputed (using the median for continuous variables and mode for categorical variables). Numbers missing on covariates are reported in the supplement. Variables were measured at the baseline visit (occurring 5.7 years prior to the start of actigraphy recording, on average) except for diabetes (ICD10 code E11 any time prior to start of actigraphy, or self-report at baseline or visit 1), diagnosed sleep disorders (ICD10 code G47 any time prior to start of actigraphy), and actigraphy variables. Healthy Diet Score ranges from 0-5 where 5 represents a healthy diet based on meeting some of the American Heart Association dietary recommendations. Family history of HTN was based on self-reported high blood pressure of father, mother, or siblings. PRS for HTN was based on the Standard PRS, which does not use UK Biobank participants in the training set. Depression/anxiety is self-reported and positive if they report seeing a doctor for nerves, anxiety, tension, or depression, or if they answered that they’ve experienced frequent (>half of days in the past 2 weeks) depressed mood or unenthusiasm/disinterest. Physical activity is the percentage of time spent in moderate or vigorous physical activity across the actigraphy recording.

The mean (SD) of sleep duration SD and SRI were 54 (30) minutes and 79 (11), respectively, with an average sleep duration of 7.3 hours (Supplementary Figures S2 and S3 provide sleep duration SD and SRI distributions). As expected, there was an inverse association between sleep duration SD and SRI (Spearman r=-0.39; Supplementary Figure S4). The distributions of participant characteristics across categories of sleep duration SD were mostly similar, although those with more irregular sleep were more likely to be current smokers, report depressive or anxiety symptoms, have diabetes or sleep disorder diagnoses, identify as evening chronotypes, and have shorter and less efficient sleep. Similar patterns were observed across SRI quintiles (Supplementary Table S3).

There were 5,713 incident hypertension cases observed over a median follow-up of 7.9 (interquartile range: 1.1) years (rate: 137 cases per 10,000 person-years). More irregular sleep duration was associated with higher risk of incident hypertension (Table 2). In the age and sex adjusted model, those with sleep duration SD of 46-60 minutes, 61-90 minutes, and >90 minutes had HRs 10%, 12%, and 25% greater than those with ≤30 minutes. After further adjustment for family history of hypertension, socioeconomic and lifestyle factors (Model 2), these estimates attenuated but remained directionally consistent. When treated continuously, each one-SD increase in sleep duration SD (about 30 minutes) was associated with a 5% higher hazard of developing hypertension (95% CI: 1.02, 1.07). The HR in Model 2 for those with >90 minutes sleep duration SD was 1.16 (95% CI: 1.06, 1.27) when comparing to the reference of ≤90 minutes (binary classification). In a sensitivity analysis we repeated Model 2 using quintiles of sleep duration SD to aid comparison to SRI quintile-based analyses and the results were very similar to the *a priori*-defined minute-based categories (Supplementary Table S4). In exploring the potential nonlinear relationship between sleep duration SD and incident hypertension using a spline model, we found that the association was linear for the bulk of the distribution, with higher hypertension risk as sleep duration SD increased up to approximately 110 minutes, representing >90% of the sample’s distribution. Above that, the risk began to plateau and then decreased (p for nonlinearity=0.03; Figure 1).

**Figure 1.**
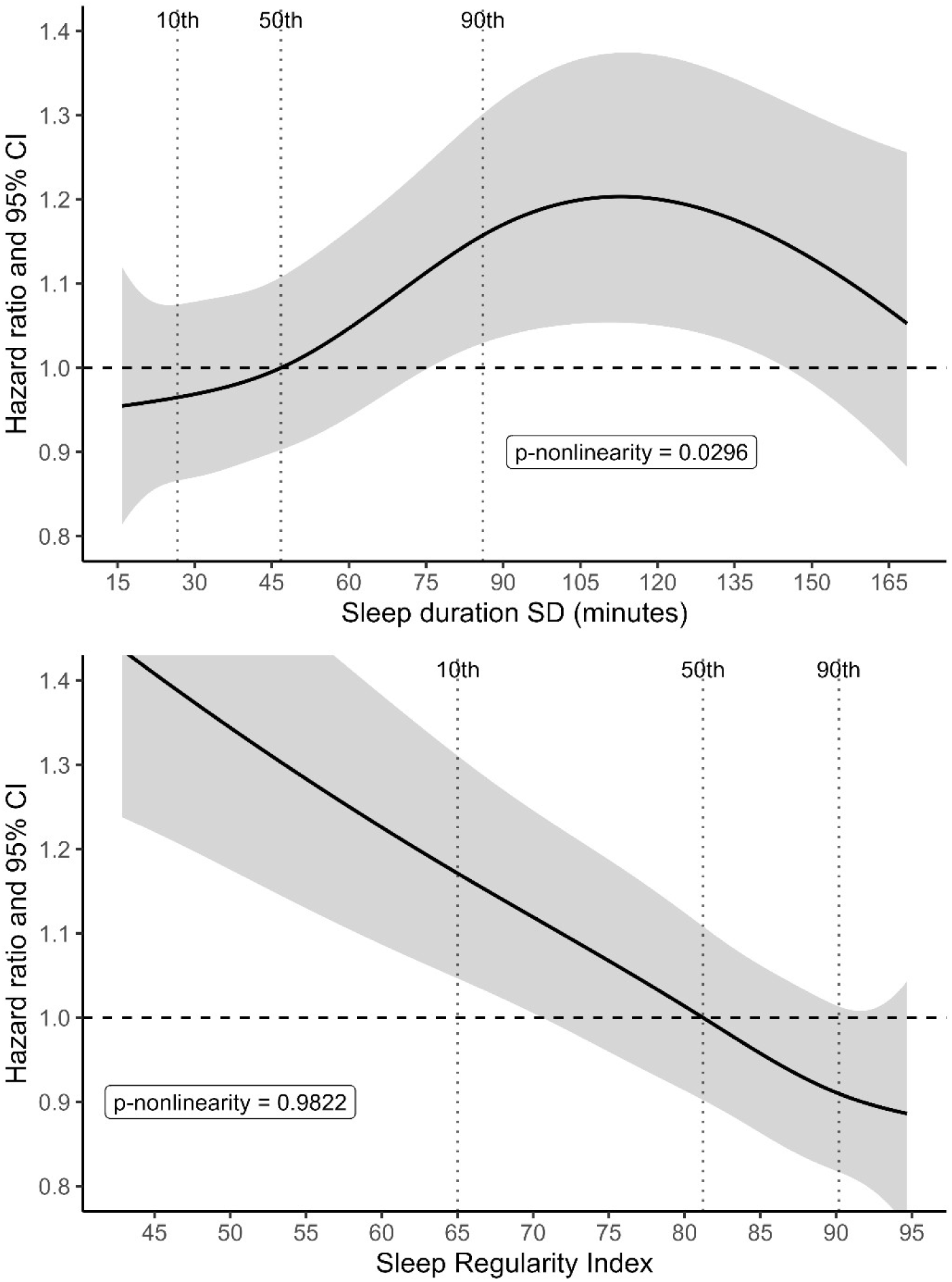
Examining potential nonlinearity in the associations between sleep regularity metrics and incident hypertension. Natural cubic spline models (forced linear at the tails) with internal knots set at the 10^th^, 50^th^ (reference value), and 90^th^ percentiles of sleep regularity metrics were used to estimate nonlinear associations with incident hypertension via Cox proportional hazards model adjusted for the following (plotted using the mean or mode values): age, sex, ethnic background (White/European, non-European), Townsend deprivation index, employment/shiftwork, family history of hypertension, smoking history (never, former, current), alcohol use (never, former, current), physical activity (proportion of time spent in moderate or vigorous activity, from actigraphy), healthy diet score. P-nonlinearity is from the likelihood ratio test comparing the model with the linear (continuous) sleep regularity term compared to the spline model. Plotted range is from the 1^st^-99^th^ percentiles. Horizontal dashed line indicates HR=1. Vertical dotted lines indicate percentile locations of internal knots.

**Table 2.** Rate, adjusted hazard ratios and 95% confidence intervals for incident hypertension by level of sleep irregularity, measured by sleep duration SD and the Sleep Regularity Index.

|  | n (cases) | person-time (yrs) | Rate<sup>a</sup> | Model 0 | Model 1 | Model 2 |
|---|---|---|---|---|---|---|
| **Sleep duration SD** |  |  |  | HR (95% CI) | HR (95% CI) | HR (95% CI) |
|---|---|---|---|---|---|---|
| Continuous, per-SD | 55,572 (5713) | 417,560 | 137 | 1.07 (1.04, 1.09) | 1.06 (1.03, 1.09) | 1.05 (1.02, 1.07) |
| Categorical |  |  |  |  |  |  |
| <= 30 min. | 8417 (897) | 62,936 | 143 | Ref. | Ref. | Ref. |
| 31-45 min. | 17,236 (1703) | 129,809 | 131 | 0.97 (0.90, 1.06) | 0.97 (0.90, 1.05) | 0.96 (0.89, 1.04) |
| 46-60 min. | 14,462 (1518) | 108,658 | 140 | 1.10 (1.01, 1.19) | 1.09 (1.00, 1.18) | 1.06 (0.98, 1.15) |
| 61-90 min. | 10,464 (1057) | 78,717 | 134 | 1.12 (1.03, 1.23) | 1.10 (1.01, 1.21) | 1.06 (0.97, 1.16) |
| > 90 min. | 4993 (538) | 37,439 | 144 | 1.25 (1.13, 1.40) | 1.22 (1.10, 1.36) | 1.18 (1.06, 1.31) |
| Binary |  |  |  |  |  |  |
| <=90 min. | 50,579 (5175) | 380,121 | 136 | Ref. | Ref. | Ref. |
| > 90 min. | 4993 (538) | 37,439 | 144 | 1.20 (1.10, 1.32) | 1.18 (1.08, 1.29) | 1.16 (1.06, 1.27) |
| **Sleep regularity index** |  |  |  |  |  |  |
| Continuous, per-SD | 55,572 (5713) | 417,560 | 137 | 0.86 (0.84, 0.88) | 0.87 (0.85, 0.89) | 0.90 (0.88, 0.92) |
| Categorical |  |  |  |  |  |  |
| Q5 | 11,891 (1052) | 90,248 | 117 | Ref. | Ref. | Ref. |
| Q4 | 11,633 (1127) | 87,775 | 128 | 1.15 (1.06, 1.25) | 1.15 (1.06, 1.25) | 1.12 (1.03, 1.22) |
| Q3 | 11,266 (1114) | 85,028 | 131 | 1.18 (1.09, 1.29) | 1.17 (1.08, 1.28) | 1.14 (1.04, 1.24) |
| Q2 | 10,832 (1137) | 81,272 | 140 | 1.25 (1.15, 1.36) | 1.22 (1.13, 1.33) | 1.16 (1.07, 1.26) |
| Q1 | 9950 (1283) | 73,238 | 175 | 1.57 (1.45, 1.71) | 1.52 (1.40, 1.65) | 1.40 (1.29, 1.52) |
| Binary |  |  |  |  |  |  |
| Q2-Q5 | 45,622 (4430) | 344,322 | 129 | Ref. | Ref. | Ref. |
| Q1 | 9950 (1283) | 73,238 | 175 | 1.38 (1.29, 1.47) | 1.34 (1.26, 1.43) | 1.26 (1.19, 1.35) |

Q: Quintile. One SD of sleep duration SD is 30.2 minutes. One SD of SRI is 10.8 points on the original SRI scale. <sup>a.</sup> per 10,000 person-years. **Model 0**: age, sex. **Model 1**: age, sex, ethnic background (White/European, non-European), Townsend deprivation index, employment/shiftwork (employed-nonshift, employed-shift, retired or not employed), family history of hypertension (based on reported illnesses of father, mother, and siblings). **Model 2**: model 1 +, smoking history (never, former, current), alcohol history (never, former, current), physical activity (proportion of time spent in moderate or vigorous activity, from actigraphy), healthy diet score.

Irregular day-to-day sleep patterns, represented by low SRI, were consistently associated with higher risk of incident hypertension (Table 2). Those in the lowest quintile (most irregular) had a 57% higher hazard of developing hypertension over the follow-up compared to those in the most regular quintile in the age- and sex-adjusted model. The HR for the same comparison was 1.40 (95% CI: 1.29, 1.52) in the fully adjusted Model 2. While there was a large comparative difference in risk from the 2^nd^ (HR=1.16) to the 1^st^ SRI quintile (HR=1.40), we observed steady risk increases across quintiles 4, 3 and 2 compared to the most regular quintile 5, with the HRs (95% CIs) of 1.12 (1.03, 1.22), 1.14 (1.04, 1.24), and 1.16 (1.07, 1.26), respectively. When treated continuously, each one-SD increase in SRI (∼11 points) towards more regular patterns was associated with a 10% lower risk of hypertension (HR [95% CI]: 0.90 [0.88, 0.92]). When treated as a binary variable, those in quintile 1 of SRI had a 26% higher risk (95% CI: 1.19, 1.35) compared to the higher 4 quintiles. The linear, dose-response association was reflected in the plotted spline model which showed consistently lower risk across increasing SRI (Figure 1; p for nonlinearity=0.98).

Across sensitivity analyses, the results remained consistent. There was mild attenuation of HRs when we further controlled for traditional cardiometabolic risk factors and comorbidities, which we hypothesized as potential mediators or colliders, and for other sleep factors (Supplementary Table S5). There was a slight increase in strength of the HRs for the most irregular groups (sleep duration SD and SRI) when we removed individuals with high baseline blood pressure readings (Supplementary Table S6). There was also a slight increase in strength of the association for those with sleep duration SD>90 minutes when we excluded those with sleep duration SD>120 minutes.

These findings were generally consistent across subgroups (Figure 2). When estimating the HR for the most irregular groups (>90 minutes sleep duration SD or Q1 of SRI) compared to the remaining more regular sample (binary classification), all subgroup estimates but one indicated higher risk for hypertension and fell in the range of HRs observed in the main analysis. The exception was among those reporting the most sleepiness at baseline (HR (95% CI) for those in quintile 1 of SRI compared to quintiles 2-5: 0.93 (0.64, 1.36)). There was no evidence of interaction except by BMI where the HR for hypertension in those with irregular sleep duration SD was higher among those with BMI≥30 than those with BMI<30 (interaction p=0.02). However, the subgroup difference by BMI was not observed for SRI.

**Figure 2.**
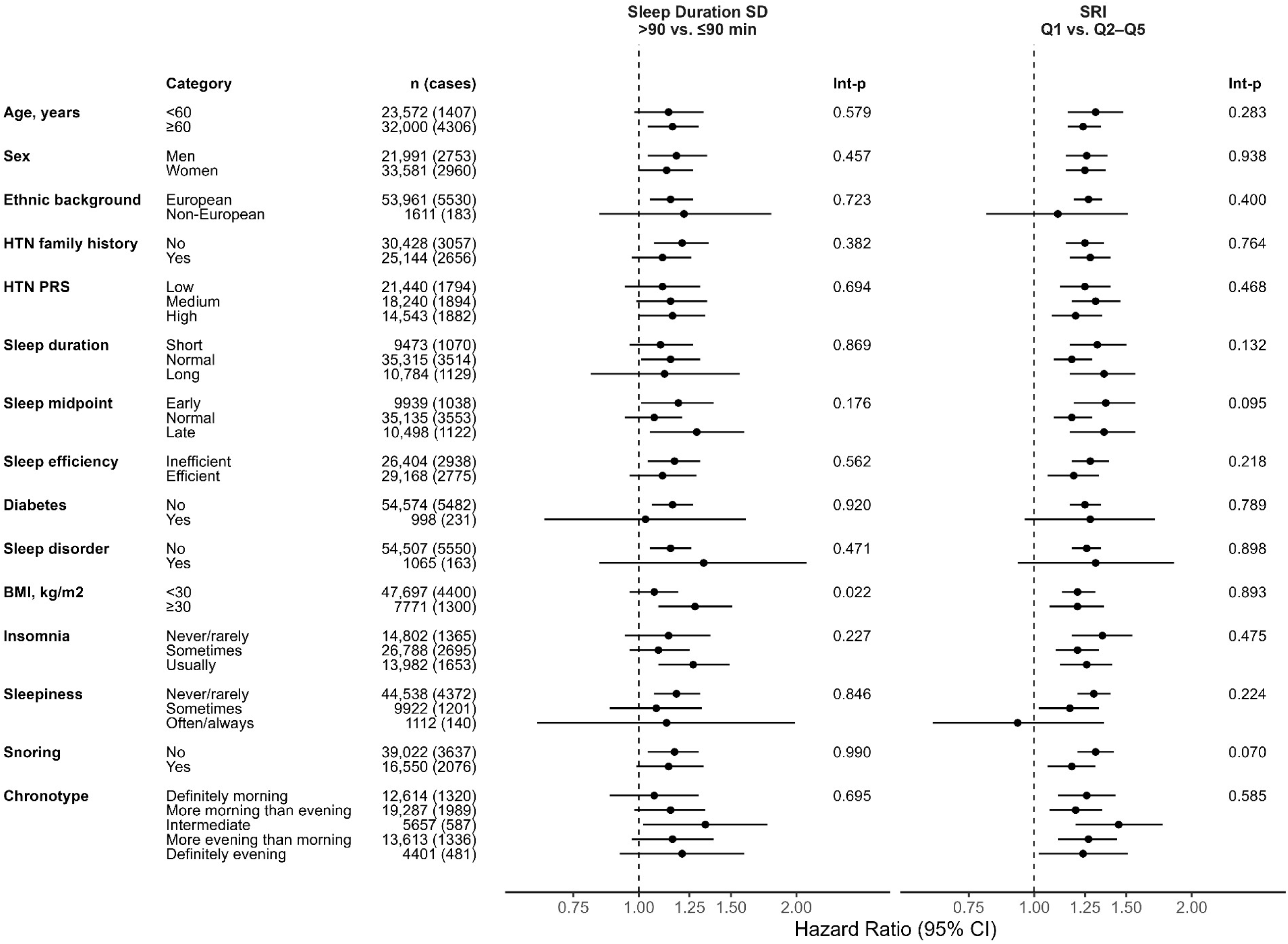
Stratified analyses: adjusted hazard ratios and 95% confidence intervals for incident hypertension by level of sleep regularity stratified by demographic, genetic, health, and sleep factors. Estimates from Cox proportional hazards models adjusted for the following: age, sex, ethnic background (White/European, non-European), Townsend deprivation index, employment/shiftwork, family history of hypertension, smoking history (never, former, current), alcohol use (never, former, current), physical activity (proportion of time spent in moderate or vigorous activity, from actigraphy), healthy diet score. Interaction p-values are from the Wald tests for the product terms included in the fully adjusted model (p-value from type-3 test for modifier variables with >2 categories). BMI: body mass index, kg/m^2^; HTN: hypertension; PRS: polygenic risk score; CI: confidence interval. Family history of HTN was based on self-reported high blood pressure of father, mother, or siblings. PRS for HTN was based on the Standard PRS, which does not use UK Biobank participants in the training set. PRS was categorized as low risk: tertile 1, medium risk: tertile 2, and high risk: tertile 3. Sleep duration, sleep midpoint (time), and sleep efficiency are estimated from the accelerometry data. These were categorized as follows: sleep duration (short: <20^th^ percentile, normal: 20^th^ to <80^th^ percentile, long: ≥80^th^ percentile), sleep midpoint (early: <20^th^ percentile, normal: 20^th^ to <80^th^ percentile, late: ≥80^th^ percentile), sleep efficiency (inefficient: <0.8, efficient: ≥0.8), Diabetes based on ICD10 code E11 any time prior to start of actigraphy, or self-report at baseline or visit 1. Sleep disorders based on ICD10 code G47 any time prior to start of actigraphy. BMI, insomnia, sleepiness, and chronotype were all measured as the UK Biobank baseline visit which occurred 5.7 years prior to the start of actigraphy recording, on average. Insomnia symptoms measured by the touchscreen question “Do you have trouble falling asleep at night or do you wake up in the middle of the night?” Sleepiness assessed by the touchscreen question “How likely are you to doze off or fall asleep during the daytime when you don’t mean to? (e.g. when working, reading or driving).” Snoring assessed from the touchscreen question “Has your snoring ever bothered other people?” Chronotype measured by the touchscreen question “Do you consider yourself to be?” with the response options shown in the figure, except that those who answered “do not know” are treated as having an intermediate chronotype, in alignment with previous research.

We found no clear evidence of multiplicative or additive interaction between sleep regularity and genetic susceptibility to hypertension (Figure 3, Supplementary Table S8). Irregular sleep was associated with higher hypertension risk within both low/medium and high genetic susceptibility strata. In joint analyses, hypertension risk was highest among participants with both irregular sleep and high genetic susceptibility (HRs of 1.80 for sleep duration SD and 1.92 for SRI). Notably, participants with high genetic susceptibility but regular sleep had higher hypertension risk than those with low/medium genetic susceptibility but irregular sleep. Similar patterns were observed when using family history of hypertension instead of PRS (Supplementary Table S9).

**Figure 3.**
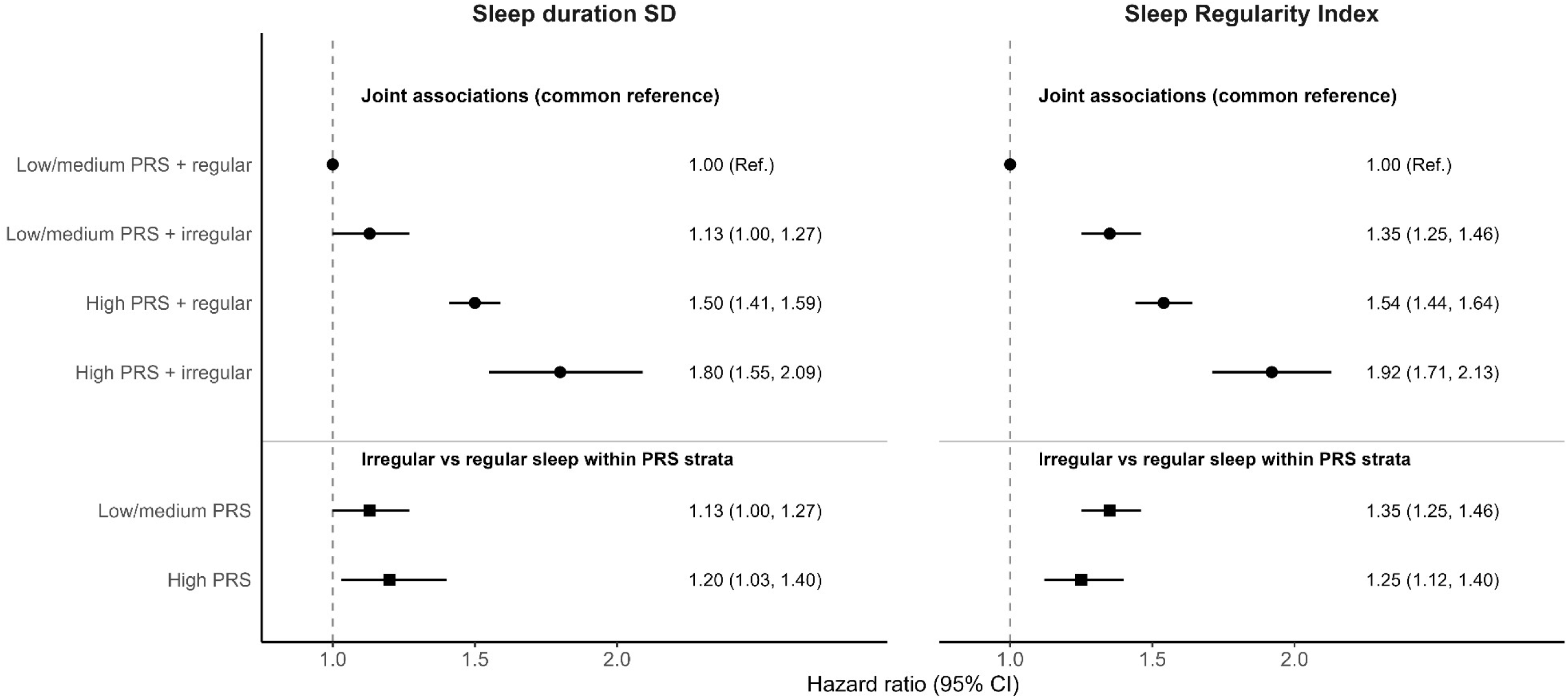
Joint associations of sleep regularity and genetic susceptibility with incident hypertension. Joint associations of sleep regularity and genetic susceptibility to hypertension with incident hypertension are presented for (A) night-to-night variability in sleep duration, measured by sleep duration standard deviation (SD), and (B) consistency of sleep-wake patterns, measured by the Sleep Regularity Index (SRI). Joint hazard ratios (HRs) and 95% confidence intervals (CIs) are presented relative to participants with low or medium genetic susceptibility and regular sleep. Associations of irregular versus regular sleep within each genetic susceptibility stratum are also presented. High genetic susceptibility was defined as the highest tertile of the hypertension polygenic risk score (PRS), and low or medium susceptibility as the lower two tertiles. Irregular sleep was defined as sleep duration SD ≥90 minutes or SRI in the lowest quintile, respectively. Models were adjusted for age, sex, the first 10 genetic principal components, Townsend deprivation index, employment and shift work, family history of hypertension, smoking, alcohol consumption, physical activity, and diet quality. Sample limited to White/European sample with genetic data, n= 52,514, cases= 5378. The PRS is the Standard PRS that did not include UK Biobank data in the training set. *P*-values for multiplicative interaction between continuous sleep regularity measures and continuous hypertension PRS were 0.161 for sleep duration SD and 0.868 for SRI. Estimates of additive and multiplicative interaction using categorical exposures are presented in Supplementary Table S8.

## Discussion

In this prospective cohort study among over 55,000 individuals, objectively measured irregular sleep patterns were consistently associated with increased risk of incident hypertension over nearly 8 years of follow-up. We observed associations across two complementary measures of sleep regularity, and these were generally stronger for the SRI than for sleep duration SD, although both metrics identified individuals at increased risk for hypertension. We observed that the most irregular sleepers had an 18 to 40% increase in hypertension risk compared to the most regular sleepers. The magnitude of these associations is comparable to other established risk factors for hypertension, such as habitual exposure to secondhand smoke and increased abdominal adiposity^35,36^. Findings were robust to comprehensive adjustment for potential confounders, additional adjustment for health and sleep- related factors, a range of sensitivity analyses, and were broadly consistent across demographic, clinical, and sleep-related subgroups. Sleep regularity and genetic susceptibility appeared to largely contribute independently to hypertension risk, where irregular sleep was associated with higher risk across levels of inherited risk. Together, these findings support sleep regularity as a potentially important and modifiable independent behavioral target for hypertension prevention in populations.

Our findings help clarify a growing but previously inconsistent literature linking irregular sleep to hypertension. Previous studies using actigraphy-derived sleep regularity have been mixed. Sleep duration SD was not associated with hypertension in the Multi-Ethnic Study of Atherosclerosis (MESA) or the Swiss CoLaus cohort^37,38^, whereas the ELSA-Brasil cohort observed modest positive associations with blood pressure and hypertension^14^. In contrast, lower SRI was consistently associated with higher blood pressure in both MESA and the Coronary Artery Risk Development in Young Adults (CARDIA) cohort^39,40^. Differences in the sociodemographic and health characteristics of study populations or sleep regularity assessment methods may partly explain these mixed findings^13,14^, and limited power may have contributed to some of the previous null findings. More recent studies have assessed sleep regularity over substantially longer periods. In the All of Us cohort, each one-hour increase in sleep duration SD measured over a median of 4.5 years using Fitbit devices was associated with a 56% increased risk of incident hypertension^41^. Similar findings were reported in a study of 12,287 individuals using under- mattress sensors over approximately 9 months^42^. Although those two studies have limited generalizability given the samples included select participants using consumer wearables or nearables that also agreed to share their data for research purposes, and the latter was unable to account for many possible confounders. Together with the prior literature, our findings strengthen the prospective evidence linking irregular sleep to incident hypertension and extend it by demonstrating consistent associations across diverse subgroups using two complementary objective measures of sleep regularity. These findings are especially relevant alongside preliminary intervention evidence that suggests sleep regularity may be a modifiable target for blood pressure reduction. In a proof-of-concept study among 11 adults with hypertension and obesity, a two-week sleep regularization intervention led to clinically meaningful reductions in blood pressure despite minimal changes in other sleep characteristics^43^.

Several biological and behavioral mechanisms may explain these associations. Blood pressure follows a strong circadian pattern, characterized by a physiological decline during sleep (nocturnal dipping) and a surge before awakening. Circadian misalignment, whereby sleep-wake cycles or other behavioral rhythms fall out of sync with endogenous circadian rhythms, may disrupt this regulation. In a controlled laboratory study, experimentally induced circadian misalignment over 10 days increased mean arterial blood pressure^44^. Irregular sleep patterns may similarly alter nocturnal dipping or the timing of the morning blood pressure surge, potentially through effects on autonomic nervous system activity and other circadian-controlled physiological processes^12^. Consistent with this hypothesis, stronger and more regular rest-activity rhythms have been associated with lower nocturnal blood pressure in young adults^45^. Greater sleep duration variability has also been associated with impaired microvascular function, suggesting vascular dysfunction as another potential pathway^46^. Additionally, irregular sleep schedules may contribute to poorer dietary behaviors, including higher energy intake and less healthy food choices^47,48^, which may subsequently contribute to elevated blood pressure.

The stronger associations observed for SRI in our study, consistent with findings from previous studies, may reflect its more comprehensive characterization of sleep-wake regularity, capturing variability in sleep timing and duration, daytime sleep-wake patterns, and nighttime awakenings through comparisons of sleep-wake states at corresponding times across consecutive days^25^. Together, these feature and the assessment of regularity on a 24-hour timescale may make SRI a more sensitive marker of behavioral irregularity than sleep duration SD, although the latter offers the advantage of more straightforward clinical interpretation.

This study has several strengths, including its prospective design; assessment of two complementary sleep regularity metrics derived from research-grade accelerometry devices using validated algorithms; a large sample size with extensive follow-up; rigorous exclusion of individuals with pre-existing hypertension; detailed participant characterization enabling adjustment for potential confounders and evaluation across clinically relevant subgroups; and robustness across multiple sensitivity analyses. Several limitations should also be considered. As an observational study, causality cannot be established and residual confounding remains possible. Sleep regularity was estimated from 7- day actigraphy recordings and may not reflect habitual long-term sleep patterns^49^; however, any resulting measurement error is likely to have been largely nondifferential and therefore would be expected to attenuate the observed associations^50^. Ascertainment of incident hypertension through linked health records may have resulted in some missed or delayed diagnoses. Finally, the predominantly White, healthier-than-average UKB sample limits the generalizability of these findings. The healthy volunteer effect may also have introduced selection bias, although its likely the direction and magnitude are difficult to predict.

## Conclusion

In this prospective study of more than 50,000 UK adults, greater sleep duration variability and more irregular sleep-wake patterns assessed via 7-day actigraphy were associated with higher risk of incident hypertension over nearly 8 years of follow-up. While these findings require replication in more diverse populations and using longer-term assessments of sleep regularity, they support hypertension as a potential pathway linking irregular sleep with CVD^5,28^. Randomized controlled trials are now needed to assess whether interventions that promote more regular sleep patterns can reduce blood pressure and hypertension risk, and thereby establish whether sleep regularity represents a causal and modifiable target for CVD prevention.

## Data Availability

The data used here are available from the UK Biobank upon approval of a data access application.

## Acknowledgments

This research has been conducted using the UK Biobank Resource under Application Number 85501. This work uses data provided by patients and collected by the NHS as part of their care and support.

## Supplemental Material

Figure S1-S4. Tables S1-S9.

